# Genetic polymorphism and evidence of signatures of selection in the *Plasmodium falciparum* circumsporozoite protein gene in Tanzanian regions with different malaria endemicity

**DOI:** 10.1101/2024.01.23.24301587

**Authors:** Beatus M. Lyimo, Catherine Bakari, Zachary R. Popkin-Hall, David J. Giesbrecht, Misago D. Seth, Dativa Pereus, Ramadhan Moshi, Ruth Boniface, Celine I. Mandara, Rashid Madebe, Jonathan J. Juliano, Jeffrey A. Bailey, Deus S. Ishengoma

## Abstract

**Background:** In 2021 and 2023, the World Health Organization approved RTS,S/AS01 and R21/Matrix M malaria vaccines, respectively, for routine immunization of children in African countries with moderate to high transmission. These vaccines are made of *Plasmodium falciparum* circumsporozoite protein (*Pfcsp)* but polymorphisms in this gene raises concerns regarding strain-specific responses and the long-term efficacy of these vaccines. This study assessed the *Pfcsp* genetic diversity, population structure and signatures of selection among parasites from areas of different malaria transmission in mainland Tanzania, to generate baseline data before the introduction of the malaria vaccines in the country.

**Methods:** The analysis involved 589 whole genome sequences generated by and as part of the MalariaGEN Community Project. The samples were collected between 2013 and January 2015 from five regions of mainland Tanzania: Morogoro and Tanga (Muheza) (moderate transmission areas), and Kagera (Muleba), Lindi (Nachingwea), and Kigoma (Ujiji) (high transmission areas). Wright’s inbreeding coefficient (F_ws_), Wright’s fixation index (F_ST_), principal component analysis, nucleotide diversity, and Tajima’s D were used to assess within-host parasite diversity, population structure and natural selection.

**Results:** Based on F_ws_ (< 0.95), there was high polyclonality (ranged from 69.23% in Nachingwea to 56.9% in Muheza). No population structure was detected in the *Pfcsp* gene in the five regions (mean F_ST_= 0.0068). The average nucleotide diversity (*π*), nucleotide differentiation (K) and haplotype diversity (Hd) in the five regions were 4.19, 0.973 and 0.0035, respectively. The C-terminal region of *Pfcsp* showed high nucleotide diversity at Th2R and Th3R regions. Positive values for the Tajima’s D were observed in the Th2R and Th3R regions consistent with balancing selection. The *Pfcsp* C-terminal sequences had 50 different haplotypes (H_1 to H_50) and only 2% of sequences matched the 3D7 strain haplotype (H_50).

**Conclusions:** The findings demonstrate high diversity of the *Pfcsp* gene with limited population differentiation. The *Pfcsp* gene showed positive Tajima’s D values for parasite populations, consistent with balancing selection for variants within Th2R and Th3R regions. This data is consistent with other studies conducted across Africa and worldwide, which demonstrate low 3D7 haplotypes and little population structure. Therefore, additional research is warranted, incorporating other regions and more recent data to comprehensively assess trends in genetic diversity within this important gene. Such insights will inform the choice of alleles to be included in the future vaccines

## Background

Malaria remains a major public health problem worldwide, with nearly half of the world’s population at risk and lives in areas where malaria transmission occurs, but with the majority of cases and deaths from Sub-Saharan (SSA) (1). In 2022, over 95% (233 million) of malaria cases and 96% of all deaths were reported from 29 countries in the World Health Organization (WHO) region of Africa (WHO – AFRO). The majority of the deaths occurred in Nigeria (31%), the Democratic Republic of the Congo (12%), Niger (6%) and the United Republic of Tanzania (4%) (2). In Tanzania, malaria is still a leading cause of morbidity and mortality mainly among children and pregnant women, and about 93% of the population lives in malaria-transmission areas (3). However, in recent years, the pattern of malaria in Tanzania has become heterogeneous, with a consistently higher burden in north-western, western and southern regions where high prevalence in school children (reaching 82%) has been reported, while other regions have consistently low to very low prevalence over the past 20 years (3).

The WHO Global Technical Strategy for Malaria 2016-2030 aims to reduce 90% of both malaria incidences and mortality rates by 90% of the 2015 levels by 2030 and proceed to the elimination targets (4). Malaria eradication which is projected by 2050 (5), will greatly depend on the use of new and innovative tools as well as enhanced use of current interventions including surveillance, monitoring and evaluation (SME), vector control, and case management [through parasitological testing with rapid diagnostic tests (RDTs) and treatment with effective antimalarials (artemisinin combination therapy-ACT)] (4). However, malaria control strategies face many challenges including emerging drug and insecticide resistance and histidine-rich protein 2/3 (*Pfhrp2*/*3*) gene deletions, limited diagnostic capacity, socio-cultural hindrance and a lack of effective vaccines (4,6,7). Urgent measures are required to get malaria elimination efforts on track and progressing to the GTS targets by 2030.

Malaria parasites have a complex life cycle in both human and mosquito hosts, including an asymptomatic stage (pre-erythrocytic), followed by a symptomatic blood stage (erythrocytic) in humans and a sexual stage in the mosquito host. The parasite also exhibits stages characterized by extensive genetic and antigenic diversity and these factors which are associated with the biology of parasites and vectors may present obstacles to anti-malarial control measures (8,9). As with other pathogens, vaccines are among the most successful and cost-effective interventions in the history of public health (10). For effective malaria control and elimination strategies, it is important to develop a malaria vaccine that provides durable protection against clinical malaria and prevents infection. Efforts to develop a malaria vaccine have been ongoing since the 1980s and focused on vaccines targeting specific stages of the parasite life cycle such as pre-erythrocytic, erythrocytic and sexual stage vaccine candidates (11–13). Antigens expressed in the pre-erythrocytic (sporozoites and liver) stages represent the ideal vaccine candidate to block the progression to clinical malaria through vaccine-based interventions (14,15).

The RTS,S/A01 is a pre-erythrocytic vaccine based on circumsporozoite protein (*Pfcsp* of the 3D7 laboratory strain of *P. falciparum*) (9). It is the first malaria vaccine to be endorsed by the WHO for routine immunization of children in moderate to high transmission areas (16). The vaccine is made of a fragment of CSP antigen fused with Hepatitis B surface antigen and AS01 adjuvant (17). It targets specific immunogenic epitopes expressed on the surface of the *Pfcsp* gene and offers better protection against malaria infections (17,18). Clinical trials of the RTS,S vaccine which were conducted in African countries including phase III trials involving children aged 5-17 months and 6-12 weeks from Mozambique, Gabon, Gambia, Ghana, Kenya and Tanzania showed significant protection against natural falciparum infections (19–23). The overall efficacy of the vaccine administered at 0-, 1-, 2-month followed by a fourth dose administered at 18 months was 36.3% (95% CI: 31.8–40.5) after an average of 48 months of follow-up (19–23).

The R21/Matrix-M vaccine is a next-generation pre-erythrocytic vaccine based on CSP, similar to the RTS,S/AS01 vaccine (24). The vaccine was created by combining the C-terminal segment of CSP from the *P. falciparum* strain NF54 (24,25). The utilized CSP portion encompasses 19 NANP repeats sourced from the central repeat region, recognized as a protective B cell epitope, along with the C-terminal region housing essential T cell epitopes. It is the second malaria vaccine endorsed by the WHO, succeeding the RTS,S/AS01 vaccine, and is recommended for use in children residing in regions with low to moderate malaria prevalence (2). This vaccine has successfully completed phase III clinical trials, involving children aged 5-36 months, at five sites in four countries across West Africa (Burkina Faso and Mali) and East Africa (Kenya and Tanzania), each with varying malaria transmission patterns. Results from this study have demonstrated a high efficacy of more than 75% against clinical malaria in African children, addressing both seasonal and perennial transmission settings (26,27). However, a study from elsewhere has indicated that the CSP-based vaccine demonstrates improved efficacy against clinical malaria when infections match the 3D7 vaccine construct at epitope haplotypes and amino acid positions. This implies that the vaccine’s efficacy may partially depend on the prevalence of the C-allele in a given geographical location (28,29).

PfCSP is predominantly a surface protein of sporozoite and plays a critical role in the invasion of hepatocytes (liver cells) (30,31). The gene encoding this protein is located on chromosome 3 of *P. falciparum* within regions that encode epitopes recognized by the human immune system. The C-terminal repeat region of the *Pfcsp* gene, which codes for epitopes recognized by anti-CSP antibodies, contains tetrameric repeats that vary in both sequence and number of tetramers (32–34). For cell-mediated immunity, the RTS,S/AS01 vaccine includes a fragment of the central NANP-NVDP repeats polymorphic B-cell epitope region and a highly polymorphic C-terminal non-repeat epitope region of *Pfcsp*, which covers CD4+ and CD8+ T-cell epitopes denoted as TH2R and TH3R, respectively (9,35). Previous studies revealed high polymorphisms in these regions within the C-terminal of the *Pfcsp* gene in natural parasite populations, which might have resulted from natural selection by the host immune system (36,37). Reports from previous studies of the population structure of *P. falciparum* genes coding for vaccine antigens, including the *Pfcsp* showed variable levels of diversity and geographic restriction of specific subgroups which may have an impact on the efficacy of malaria vaccines based on this gene in specific geographic regions (36). In particular, comparative diversity studies of the 3D7 *Pfcsp* gene showed that only 0.2% to 5.0% of the vaccine strains matched the global *Pfcsp* gene (38). A larger number of these studies described polymorphisms in the *Pfcsp* gene at the global level, with limited local-specific studies.

Therefore, there is an urgent need to explore the extent of genetic diversity and natural selection of the malaria vaccine antigen PfCSP in natural parasite populations in Tanzania and other countries where the RTS,S/AS01 and R21/Matrix-M vaccines will be deployed in the near future. This study investigated the genetic diversity and population structure of the *Pfcsp* gene within five regions of Tanzania with varying malaria transmission intensities. In addition, the study aimed to determine whether genetic diversity and signatures of selection in this important gene are present. The findings present baseline data before the Ministry of Health rolls out any of the malaria vaccines in an effort to eliminate malaria in Tanzania.

## Materials and methods

### Study area

Field studies that generated samples and data for this study were conducted between May 2013 and January 2015 as part of the Pathogen Diversity Network Africa (PDNA) baseline surveys in 15 countries from Sub-Saharan Africa (SSA) (39–41). In Tanzania, the studies covered five districts from five regions with varying malaria transmission that are located more than 800 km apart. The study districts included Muheza in the Tanga region and Morogoro (moderate to low transmission area), Ujiji in Kigoma, Muleba in Kagera and Nachingwea in Lindi (all with high transmission), as described elsewhere (39) (**Figure 1**).

**Figure 1:**
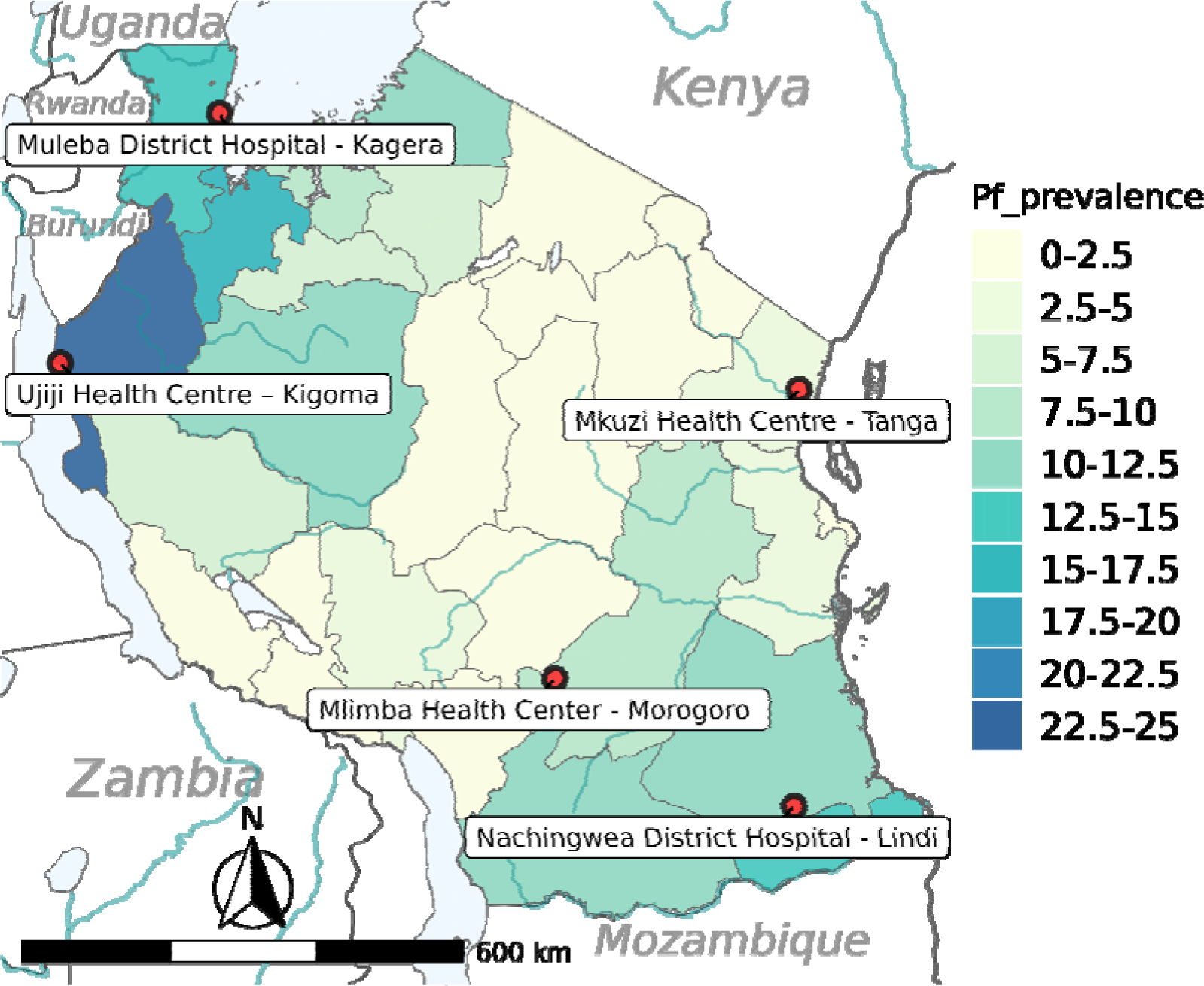
Map of Tanzania showing *P. falciparum* prevalence as a color gradient, and the location of the study regions (75).

### Study design and sampling

The samples were collected through a cross-sectional study (CSS), which was conducted in five districts of Morogoro, Muheza, Muleba, Nachingwea and Ujiji in 2013 while other samples were collected from two therapeutic efficacy studies (TES) which were conducted at Mkuzi Health Centre in 2013, and in Muheza designated district hospital and Ujiji health centre from May 2014 to January 2015 as described elsewhere (39). The TES were conducted according to the WHO protocol as previously described (42,43).

### Sample collection, processing, DNA extraction and Sequencing

Whole blood samples were collected from patients with malaria infections who were initially screened with rapid diagnostic tests for malaria (mRDTs) and the infections were confirmed using microscopy as described earlier (39). Human white blood cells (WBCs) were depleted from whole blood samples using CF11 cellulose columns (44), and parasite genomic DNA was extracted using QIAamp DNA blood midi kits (Qiagen GmbH, Hilden, Germany) following the manufacturer’s instructions. DNA samples were shipped to the Wellcome Trust Sanger Institute in Hinxton, UK, for whole genome sequencing (WGS) using the Illumina HiSeq platform as part of the MalariaGEN *Plasmodium falciparum* Community Project (45). Illumina sequencing libraries (200 bp insert) were aligned to the reference *P. falciparum* 3D7 genome, after which variant calling was conducted via the customized GATK pipeline (45). Each sample was genotyped for 797,000 polymorphic biallelic coding single nucleotide polymorphisms (SNPs) across the genome, ensuring a minimum of 5× paired-end reads coverage across each variant per sample. The dominant allele was retained in the genotype file at loci with mixed reads (reference/non-reference).

### Sequence acquisition and pre-processing

WGS data of 589 *P. falciparum* samples collected from the five districts of Morogoro (n = 32), Muheza (n = 297), Muleba (n = 52), Nachingwea (n = 65) and Ujiji (n = 143) were downloaded from the MalariaGEN *Plasmodium falciparum* Community (Pf7k) project in variant call format (VCF) at https://www.malariagen.net/resource/34. The VCF file was generated following standardized protocols (46). In brief, reads mapping to the human reference genome were discarded before all analyses, and the remaining reads were mapped to the *P. falciparum* 3D7 v3 reference genome using Burrows-Wheeler aligner (bwa) mem version 0.7.15 (47). Binary Alignment Map (BAM) files were created using the Picard tools (48) CleanSam, FixMateInformation and MarkDuplicates version 2.6.0 and GATK v3 (49) base quality score recalibration. Potential SNPs and indels were discovered by running GATK’s HaplotypeCaller independently across each sample BAM file and genotyping these for each of the 16 reference sequences (14 chromosomes, 1 apicoplast and 1 mitochondria) using GATK’s CombineGVCFs and GenotypeGCVFs. SNPs and indels were filtered using GATK’s Variant Quality Score Recalibration (VQSR). All variants with a variant quality score log-odds (VQSLOD) score ≤0 were filtered out and functional annotations were applied using snpEff version 4.1. Genome annotation was performed using vcftools version 0.1.10 and masked if they were outside the core genome. Only biallelic SNPs from chromosome 3 that passed all VCF filters were retained. Then SNP variants for the entire *Pfcsp* were extracted from chromosome 3 (position: 221,323–222,516) for further analysis.

### Population genetics analysis

The analysis included determining the minor allele frequency (MAF) distribution for all putative SNPs within *Pfcsp* using Plink1.9, and rare alleles (MAF ≤ 0.01) were removed from the analysis. Wright’s inbreeding coefficient (F_ws_) (50) was determined using R package moimix (51). F_ws_ refers to the number of different parasite strains contained within an infection, and it ranges from 0 to 1. A sample is classified as having multiple infections (polyclonal) when F_ws_ < 0.95 and a single infection (monoclonal) when F_ws_ ≥ 0.95. A Pearson chi-square test was used to determine the statistical significance of any differences observed in F_ws_estimates between the populations, and a standard threshold of *p* < 0.05 was considered statistically significant.

To assess gene flow between parasite populations, genetic differentiation was first estimated using the Wright Fixation index (F_ST_) (52) using Vcftools v0.1.5 and population structure was determined using principal component analysis (PCA) as implemented in PLINK1.9. F_ST_ <0.05 indicates minimal population differentiation or gene flow between population pairs. Haplotype diversity (the number of two randomly selected strains within the population having different haplotypes) in *Pfcsp* was determined by studying the variants in the C-terminal region of the gene (221422-221583 on chromosome 3). A total of 229 FASTA DNA sequences were reconstructed from the retained monoclonal VCF samples and the *Pfcsp* sequence of the 3D7 strain was included as a reference. The monoclonal VCF samples were obtained based on the F_ws_ results, where samples with F_ws_ ≥ 0.95 were considered a single infection (monoclonal). The average number of pairwise nucleotide diversity (K), haplotype diversity (Hd) and nucleotide diversity were calculated using DnaSP version 6.12.03 (53). To assess the genetic relationships between *Pfcsp* C-terminal haplotypes in the five districts, the haplotype networking for 229 sequences of *Pfcsp* was analyzed using NETWORK version 10.2 with the Median-joining algorithm (54).

Nucleotide diversity (π) was used to measure the degree of polymorphism within the parasite population, and it was analyzed by calculating the pairwise difference between all possible pairs of individuals in the samples using Vcftools v0.1.5 (55) in the sliding windows, with a size of 1000 bp. The Tajima’s D statistical test (56) was used to detect evidence of balancing selection or purifying selection in the *Pfcsp*. Tajima’s D was calculated in *Pfcsp* monoclonal samples using Vcftools v0.1.5 in 1000 bp sliding windows. Tajima’s D test compares the average pairwise differences (π) and the total number of segregating sites (S) (57) and it was assesed using MEGA software version 11.0.13 (58) and DnaSP version 6.12.03 (53).

## Results

### Within-host parasite diversity estimation and statistical analysis

Within-host diversity in the Pfcsp gene was assessed using the inbreeding coefficient (F_ws_) and theresults indicated high within-host diversity (polyclonality), varying from 69.23% in Nachingwea to 56.9% in Muheza. Nevertheless, the observed difference did not reach statistical significance (***p*= 0.3596**)(**Figure 2**)

**Figure 2:**
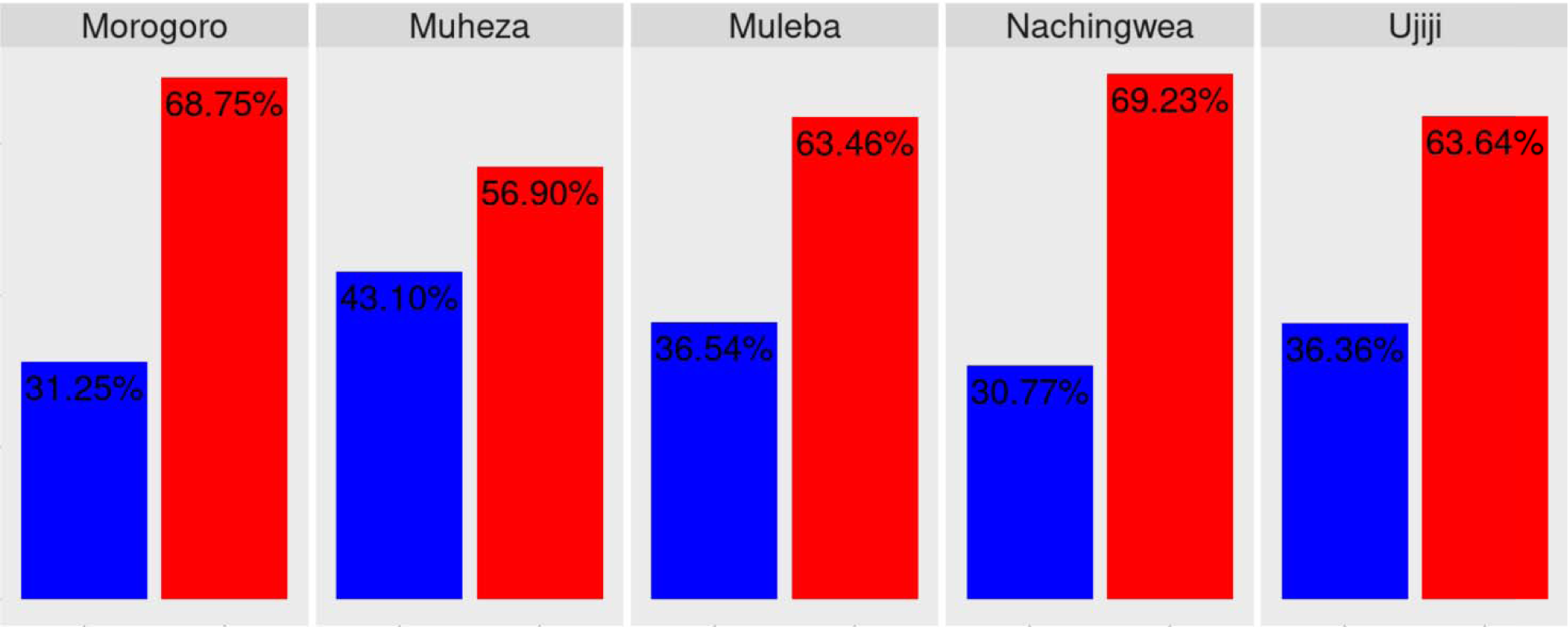
**With in-host** diversity in the samples from Morogoro, Muheza, Muleba, Nachingwea, and Ujiji populations. The differences were not statistically significant (*p*= 0.3596).

### Genetic differentiation and structure analysis

Genetic differentiation in the Pfcsp population was analyzed by calculating Wright’s fixation index (F_ST_). The mean F_ST_ across five districts—Morogoro, Muheza, Muleba, Nachingwea and Ujiji was very low at 0.0068 (which is < 0.05) (**Figure 3a**). This indicates low genetic differentiation among the parasite populations or a high exchange of genetic materials despite the districts being geographically separated with a distance of over 800 km. Principal component analysis (**Figure 3b**) showed no population structure between the five districts. The overall results suggest gene flow between the populations.

**Figure 3:**
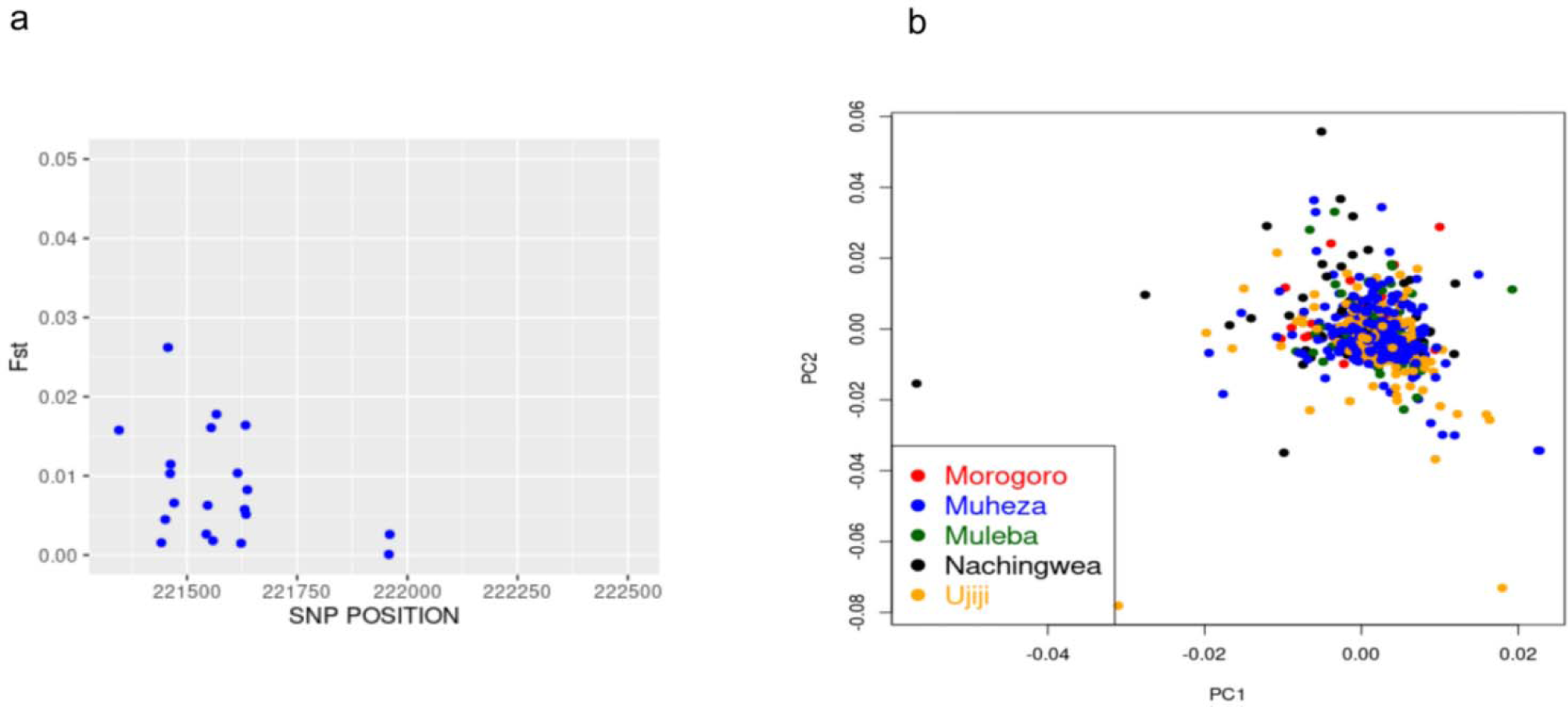
Population differentiation and structure. **a**. Weir and Cockerham’s F_ST_ among parasites from Morogoro, Muheza, Muleba, Nachingwea and Ujiji. **b**. The plot of the first (PC1) and second principal components (PC2) of samples from Morogoro, Muheza, Muleba, Nachingwea and Ujiji.

### Genetic diversity of *Pfcsp* C-terminal within parasite populations

To assess the extent of genetic diversity and similarity within and between the five populations, an investigation into the diversity in the C-terminal region of Pfcsp (221,422–221,583) was done and summarized in a median-join haplotype diversity network (**Figure 4**). In total, 50 haplotypes were observed among the 230 Pfcsp sequences, including the 3D7 sequence. Of these, more haplotypes were identified in Ujiji (24) and Muheza (27) while the other districts had fewer haplotypes between 8 and 14 (Morogoro = 8, Muleba, 14, Nachingwea = 14). The RTS,S vaccine haplotype (Pf3D7-type) was found in 2% (H_50) of all samples. Four haplotypes (8%) were shared by *Pfcsp* sequences from all five regions, indicating genetic closeness between these populations. Haplotype 5 emerged as the most prevalent *Pfcsp* C-terminal haplotype, representing 16.5% (38/230) of the isolates. Among the haplotypes, 50% appeared only once (singleton haplotypes).

**Figure 4:**
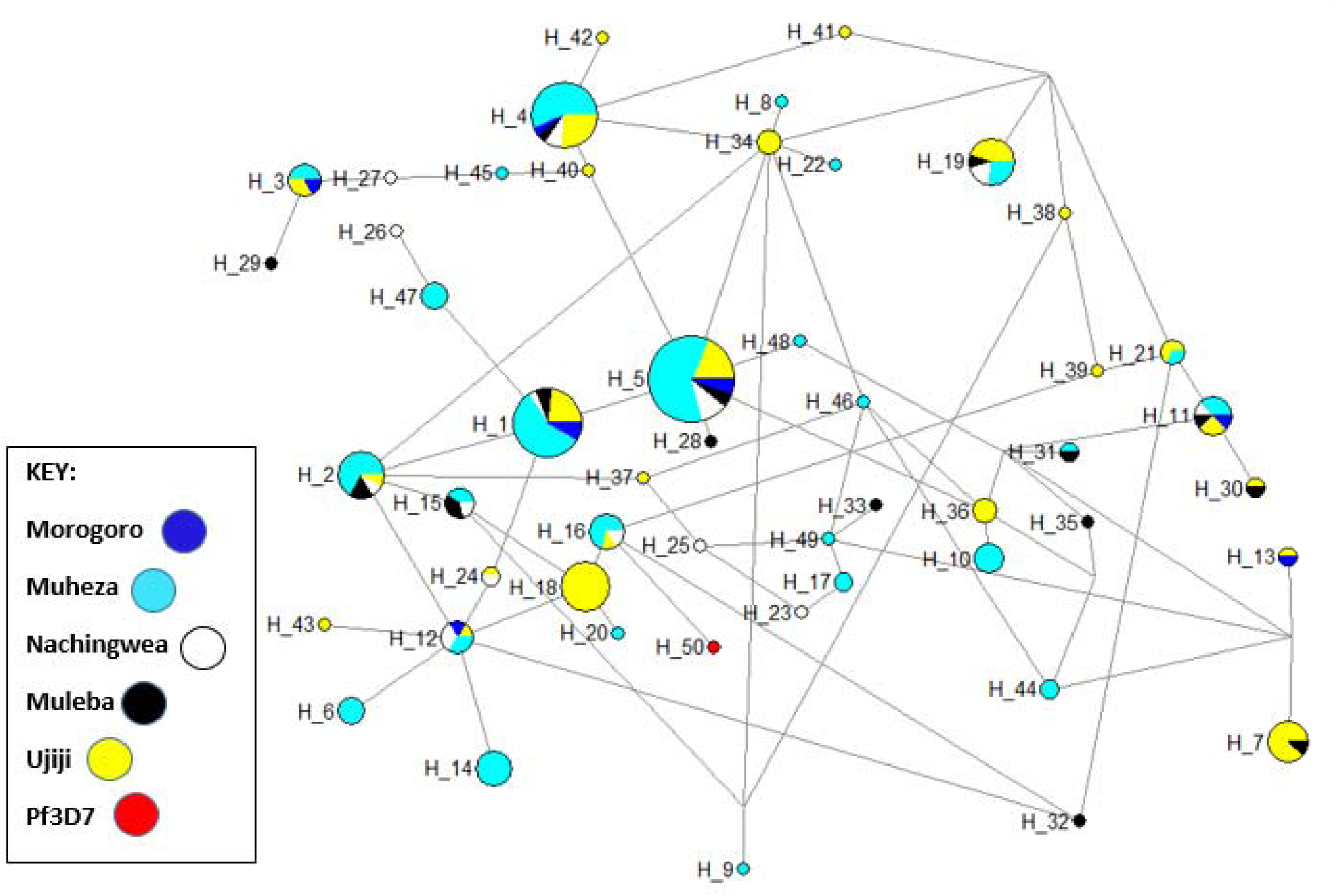
Haplotype network analysis of the *Pfcsp* gene among parasites from Morogoro, Muheza, Muleba, Nachingwea and Ujiji.The size of each node indicated the proportion of the total haplotype frequencies. Each haplotype is donated as “H” with the vaccine strain 3D7 donated as “3D7” in Hap_50. The color of each node corresponds to the site of sample origin.

### Nucleotide diversity and signatures of selection in the C-terminal non-repeat region of the *Pfcsp* gene

The average number of pairwise nucleotide differences (*K*) was higher in Muleba (4.1) and Ujiji (4.0) compared to other districts with 3.3 - 3.4 (in Morogoro = 3.4, Muheza = 3.4, and Nachingwea = 3.3). The highest haplotype diversity (Hd) was observed in Muleba (0.976±0.008) and Ujijji (0.973±0.005) and the lowest was observed in Morogoro (0.917±0.035). The highest nucleotide diversity (π) was observed in Muleba (0.0038) and Muheza (0.0037) and the lowest was observed in Morogoro (0.0029) but the differences were not significant (*p* >0.05). Evidence of natural selection in the C-terminal non-repeat region of the *PfCcsp* gene was tested using Tajima’s D and there were slightly positive values of 1.3, 1.3, 0.76 and 0.16 in Muheza, Muleba, Nachingwea and Ujiji, respectively; while Morogoro showed negative value of −0.21. However, these values suggest evidence of weak balancing in the population (**Table 1**).

**Table 1:** Diversity of the *Pfcsp* C-terminal region of samples included in the network analysis.

| Population | n | H | s | k | $\pi$ | Hd | Tajima's D |
| --- | --- | --- | --- | --- | --- | --- | --- |
| Morogoro | 10 | 8 | 15 | 3.4 | 0.0198 | 0.978±0.035 | -0.235 |
| Muheza | 128 | 27 | 18 | 4.3 | 0.0027 | 0.925±0.005 | 0.12 |
| Muleba | 19 | 14 | 15 | 4.008 | 0.0037 | 0.970±0.016 | 0.44 |
| Nachingwea | 20 | 14 | 11 | 3.920 | 0.00033 | 0.953±0.008 | 0.41 |
| Ujiji | 52 | 24 | 17 | 4.008 | 0.0027 | 0.947±0.005 | 0.16 |

*n* number of sequences, *h* number of unique haplotypes, *s* number of segregating sites, *k* average of pairwise nucleotides differences, $\pi$ nucleotide diversity, *Hd* haplotype diversity

The nucleotide diversity peaked at the Th2R and Th3R T-cell epitopes, while the connecting region between Th2R and Th3R was conserved. Nucleotide diversity showed slightly different values between populations according to their geographic origin (**Figure 5**), with Muleba showing a higher peak > 0.07 in Th2R epitopes than the other four districts. The extended haplotype homozygosity revealed some extended haplotypes from the focal SNP locus 221,554 in all populations, but no long range haplotypes extended beyond 221,554 (**Figure 6**)

**Figure 5:**
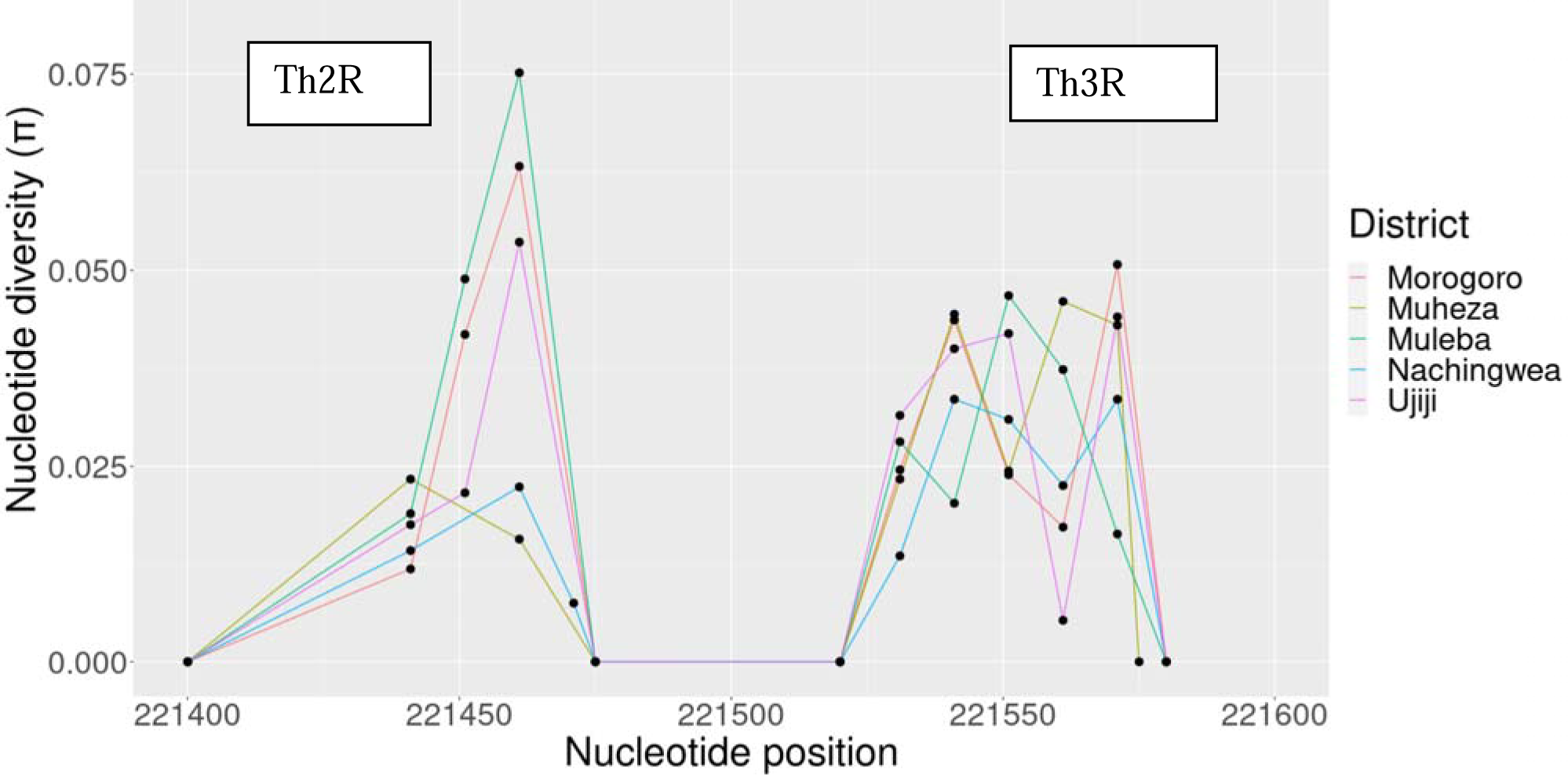
Nucleotide diversity of the C-terminal region of the *Pfcsp* gene indicated high diversity in both Th2R and Th3R, while the connecting region between Th2R and Th3R remained conserved.

**Figure 6:**
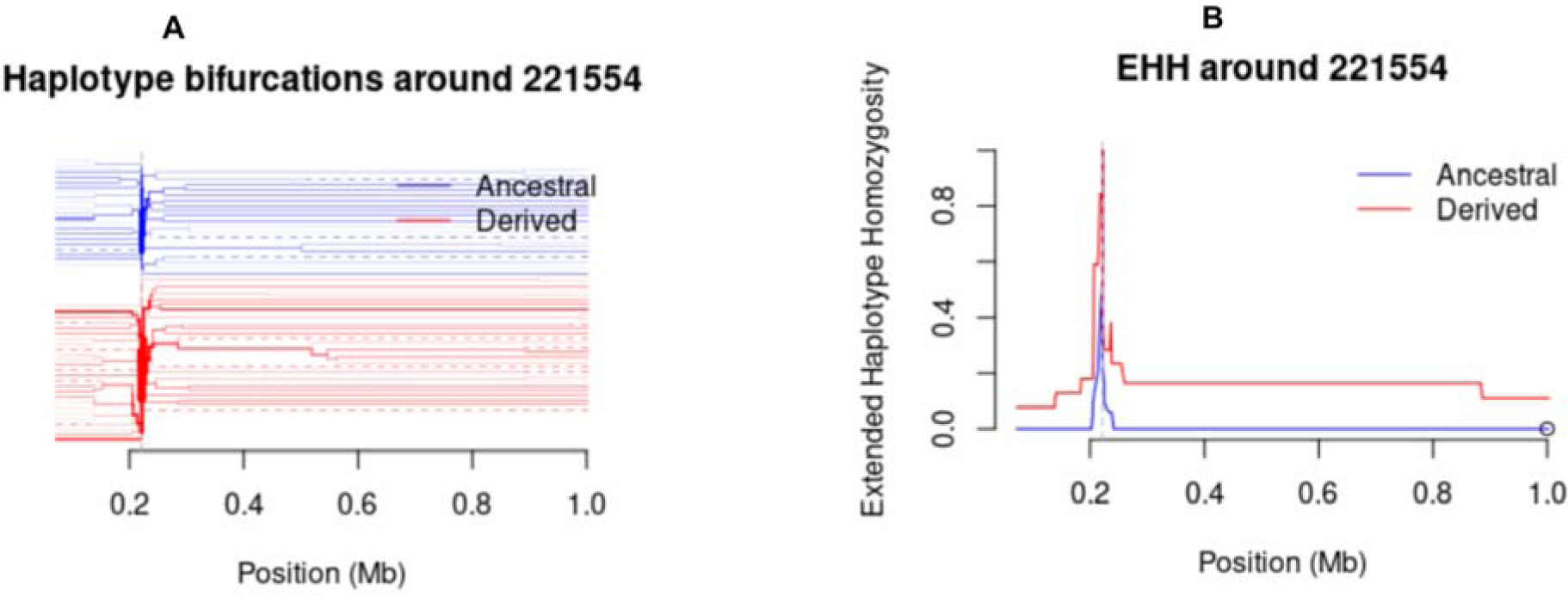
Plots show Bifurcation diagram and Extended Haplotype Homozygosity (EHH). a) Bifurcation diagrams showing the breakdown of these extended haplotypes from increasing distances in the parasite population **b)** Plots of EHH showing extended haplotypes from a focal SNP locus (221,554).

## Discussion

Tanzania is one of the malaria-endemic countries with a high number of cases and deaths in SSA and malaria is still a major health problem despite intensified interventions in the past two decades (2). Recent studies show that malaria transmission in Tanzania has become heterogeneous with persistently high burden in the western, north-west and southern regions for over 20 years (59) but lower transmission intensities in other regions attributed to the ongoing control efforts. The Tanzanian NMCP is implementing targeted interventions to reduce the burden in areas with high transmission and eliminate malaria in regions that have consistently reported low transmission over the past 20 years (60). Among the interventions, NMCP is considering the deployment and use of malaria vaccines following their approval by WHO in 2021 and 2023 (2,61). This study was therefore undertaken to generate baseline data on the genetic diversity, population structure and signature of selection in the *Pfcsp* gene before the roll-out of malaria vaccines in Tanzania. The data will potentially be useful in monitoring the performance of the vaccines after their introduction and use in Tanzania to look for shifts in variations – or new variations– that may arise due to vaccine pressure.

In general, the study showed no clear differentiation in the patterns of the *Pfcsp* gene polymorphism despite the study sites being widely separated geographically (with over 800 km apart), which is consistent with global patterns of *Pfcsp* variations which have been reported elsewhere (62,63). The lack of clear differentiation was according to the pairwise index of differentiation analysis (F_ST_ below 0.05) and confirmed by PCA. Similar findings were reported in Ghana where *pfcsp* sequences collected from two sites located over 784.4 km apart showed a largely homogeneous population with F_ST_ below 0.05, and without any population structure as detected by PCA (62). This and other studies of the *Pfcsp* gene suggested a continued gene flow among parasites or human population mixing between the areas which might have contributed to the observed findings (62,64,65). High genetic diversity and a lack of population differentiation may have important implications related to the efficacy and emergence as well spread of vaccine-resistant parasites (66,67).

The C-terminal of the *Pfcsp* gene displayed high genetic diversity in the study areas. A comparative sliding window plot analysis of nucleotide diversity in the C-terminal region displayed predictable results of two peaks at the Th2R and Th3R regions, suggesting that genetic variations were mainly concentrated in these regions. Differences were also observed among parasites from different geographical locations. The overall nucleotide diversity was 0.0035 with Muleba showing a high peak in Th2R epitopes. A similar pattern of nucleotide polymorphism was found in other African countries including Malawi, The Gambia, Nigeria, Ghana, Mali and Guinea (8,38,62).

The overall haplotype diversity in the C-terminal of the *Pfcsp* gene in Tanzanian isolates was 0.937 which is similar to those previously observed in Cameroon, The Gambia, Ghana, Senegal, Congo, Malawi and Guinea but higher than those observed in non-African countries (8,68). This suggests that the C-terminal of the *Pfcsp* gene in Africa has a higher level of diversity compared to other parts of the world (37). The values of Tajima’s D in the C-terminal region of the *Pfcsp* exhibited slightly positive values which suggest evidence of weak balancing selection in response to host immune pressure on this region (8). Similar results have been reported showing weak variant-specific selection within this region, as shown by a study in Malawi (69). Evidence of balancing selection has been reported in other malaria vaccine candidates such as the extracellular domains of *P. falciparum* apical membrane antigen (PfAMA-1), Cell-Traversal protein for Ookinetes and Sporozoites (CelTOS), Thrombospondin Related Adhesion Protein (TRAP), Liver Stage Antigen 1 (LSA1) and merozoite surface proteins (MSP1) (70–73). Evidence of recent directional selection (iHS) was not observed in all study areas and extended haplotype homozygosity showed no extended haplotypes from the focal SNP locus (221,554)(69,74).

The haplotype network analysis in the C-terminal showed a sequence connection between the five areas, which suggests they were closely related. This relatedness is in agreement with the F_ST_ and PCA results which showed limited separation in isolates from the five districts. The current RTS,S, recombinant vaccine was constructed with *pfcsp* of *P. falciparum* 3D7 strain, but in Tanzanian isolates, only 0.7% 3D7-matched Tanzanian *pfcsp* haplotypes. Low matching of the 3D7 to the natural *Pfcsp* has been reported in many other studies in Africa (8,38,64,68). This data is consistent with other studies conducted across Africa and worldwide, which demonstrate low 3D7 haplotypes and little population structure. However, other studies of the genetic diversity and protective efficacy of the RTS,S/AS01 vaccine suggested that high levels of mismatch between natural isolates and 3D7 may weaken vaccine efficacy (28,68). In addition, the high degree of location-specific *Pfcsp* diversity observed in Tanzania might result in differences in vaccine efficacy potentially reducing RTS,S/AS01 vaccine effectiveness. Monitoring of differential vaccine efficacy according to *Pfcsp* haplotypes during RTS,S/AS01 implementation programmes will be valuable, particularly in high transmission areas, where a post-vaccination expansion of non-vaccine haplotypes is likely to be observed which could lead to reduced vaccine efficacy and vaccine breakthrough infections (64). Going forward and ensuring the vaccine is efficacious in all endemic areas, the design of a malaria vaccine may consider developing a vaccine that works for specific regions based on the genetic architecture of the parasites found in those areas.

## Conclusion

These findings demonstrate a significant level of genetic diversity in the *Pfcsp* gene within the study areas. Although there is limited population differentiation and structure, there is a high degree of polyclonality, particularly in Morogoro, Muleba and Nachingwea. Furthermore, there is no genetic structure across all the study districts, indicating a high level of gene flow and genetic exchange within the *Pfcsp* gene. In addition, the C-terminal region of the *Pfcsp* gene from Muleba and Muheza exhibited higher nucleotide diversity compared to Morogoro, Nachingwea and Ujiji. Additionally, the *Pfcsp* gene displayed slight positive Tajima’s D values in all five studied *P. falciparum* populations. These findings are consistent with the hypothesis of balancing selection acting on the Th2R and Th3R regions of the gene. However, it is important to note that further studies and continuous monitoring of the *Pfcsp* gene are necessary. These future investigations should encompass other regions and incorporate more recent data to comprehensively assess the current diversity and dynamics of the *Pfcsp* gene. By expanding the understanding of the genetic variations within the *Pfcsp* gene, it creat an opportunity to improve the knowledge of the parasite’s adaptability and contribute to the development of effective malaria control strategies

## Data Availability

Data are available under the MalariaGEN terms of use for the Pf Community Project: https://www.malariagen.net/data/terms-use/p-falciparum-community-project-terms-use.

## Availability of data and materials

The datasets generated during the current study are available in the MalariaGEN *Plasmodium falciparum* Community (Pf7k) Project https://www.malariagen.net/resource/34.

## Acknowledgement

Authors would like to thank the study participants, local health workers and technicians who participated in the sample collection. Whole genome sequencing was done at the Wellcome Sanger Institute as part of the MalariaGen *Plasmodium falciparum* Community Project.

## Ethical approval

The study utilized sequence data downloaded from the open-source, MalariaGEN (https://www.ncbi.nlm.nih.gov/pmc/articles/PMC2928508/). The study protocols for the projects that collected the samples were reviewed and approved by the Tanzanian Medical Research Coordinating Committee (MRCC) of the National Institute for Medical Research (NIMR). Permission to conduct the study in Morogoro, Muheza – Tanga, Muleba – Kagera, Nachingwea - Lindi and Ujiji - Kigoma was sought in writing and obtained from the district and regional medical officers. Written informed consent was obtained from patients or parents/guardians in the case of children. Appropriate information (about the study and the protocol/methods) in a language that was understood by the parents/guardians of the study patients was compiled and provided before consent was obtained. Permission to publish was sought and obtained from the Director General of NIMR

## Authors’ contributions

DSI - formulated the original idea, supervised data analysis and wrote the manuscript

BL - performed the analysis and wrote the manuscript

CB, ZPH and DG - supported the data analysis, reviewed and edited the manuscript

RB, MS and CIM - conceived the idea, implemented the field surveys, reviewed and edited the manuscript

DP and RM - reviewed and edited the manuscript

JJ, JB, and DSI - critically reviewed the manuscript

All authors contributed to the article and approved the submitted version.

## Bibliography

1. WHO. World malaria report 2021. WHO; 2021.

2. WHO. World Malaria Report 2022. WHO [Internet]. 2022 [cited 2022 Dec 10]; Available from: https://apps.who.int/iris/rest/bitstreams/1484818/retrieve

3. Mitchell CL, Ngasala B, Janko MM, Chacky F, Edwards JK, Pence BW, et al. Evaluating malaria prevalence and land cover across varying transmission intensity in Tanzania using a cross-sectional survey of school-aged children. Malar J. 2022 Mar 9;21(1):80.

4. WHO. GLOBAL TECHNICAL STRATEGY FOR MALARIA. WHO. 2021;

5. Feachem RGA, Chen I, Akbari O, Bertozzi-Villa A, Bhatt S, Binka F, et al. Malaria eradication within a generation: ambitious, achievable, and necessary. Lancet. 2019 Sep 21;394(10203):1056–112.

6. Dhiman S. Are malaria elimination efforts on right track? An analysis of gains achieved and challenges ahead. Infect Dis Poverty. 2019 Feb 13;8(1):14.

7. Guyant P, Corbel V, Guérin PJ, Lautissier A, Nosten F, Boyer S, et al. Past and new challenges for malaria control and elimination: the role of operational research for innovation in designing interventions. Malar J. 2015 Jul 17;14:279.

8. Huang H-Y, Liang X-Y, Lin L-Y, Chen J-T, Ehapo CS, Eyi UM, et al. Genetic polymorphism of Plasmodium falciparum circumsporozoite protein on Bioko Island, Equatorial Guinea and global comparative analysis. Malar J. 2020 Jul 13;19(1):245.

9. Casares S, Brumeanu T-D, Richie TL. The RTS,S malaria vaccine. Vaccine. 2010;28(4880–94).

10. Koff WC, Gust ID, Plotkin SA. Toward a human vaccines project. Nat Immunol. 2014 Jul;15(7):589–92.

11. Nadeem AY, Shehzad A, Islam SU, Al-Suhaimi EA, Lee YS. Mosquirix^TM^ RTS, S/AS01 vaccine development, immunogenicity, and efficacy. Vaccines (Basel). 2022 Apr 30;10(5).

12. Duffy PE, Patrick Gorres J. Malaria vaccines since 2000: progress, priorities, products. npj Vaccines. 2020 Jun 9;5:48.

13. Le Roch KG, Chung DWD, Ponts N. Genomics and integrated systems biology in Plasmodium falciparum: a path to malaria control and eradication. Parasite Immunol. 2012 Mar;34(2–3):50–60.

14. Mohamed NS, AbdElbagi H, Elsadig AR, Ahmed AE, Mohammed YO, Elssir LT, et al. Assessment of genetic diversity of Plasmodium falciparum circumsporozoite protein in Sudan: the RTS,S leading malaria vaccine candidate. Malar J. 2021 Nov 10;20(1):436.

15. Kurtovic L, Drew DR, Dent AE, Kazura JW, Beeson JG. Antibody Targets and Properties for Complement-Fixation Against the Circumsporozoite Protein in Malaria Immunity. Front Immunol. 2021 Dec 1;12:775659.

16. WHO. WHO Guidelines for malaria. WHO Geneva, Switzerland. 2022;

17. Stoute JA, Slaoui M, Heppner DG, Momin P, Kester KE, Desmons P, et al. A preliminary evaluation of a recombinant circumsporozoite protein vaccine against Plasmodium falciparum malaria. RTS,S Malaria Vaccine Evaluation Group. N Engl J Med. 1997 Jan 9;336(2):86–91.

18. Regules JA, Cummings JF, Ockenhouse CF. The RTS,S vaccine candidate for malaria. Expert Rev Vaccines. 2011 May;10(5):589–99.

19. Asante KP, Abdulla S, Agnandji S, Lyimo J, Vekemans J, Soulanoudjingar S, et al. Safety and efficacy of the RTS,S/AS01E candidate malaria vaccine given with expanded-programme-on-immunisation vaccines: 19 month follow-up of a randomised, open-label, phase 2 trial. Lancet Infect Dis. 2011 Oct;11(10):741–9.

20. Bojang KA, Milligan PJ, Pinder M, Vigneron L, Alloueche A, Kester KE, et al. Efficacy of RTS,S/AS02 malaria vaccine against Plasmodium falciparum infection in semi-immune adult men in The Gambia: a randomised trial. Lancet. 2001 Dec 8;358(9297):1927–34.

21. Aponte JJ, Aide P, Renom M, Mandomando I, Bassat Q, Sacarlal J, et al. Safety of the RTS,S/AS02D candidate malaria vaccine in infants living in a highly endemic area of Mozambique: a double blind randomised controlled phase I/IIb trial. Lancet. 2007 Nov 3;370(9598):1543–51.

22. RTS,S Clinical Trials Partnership. Efficacy and safety of RTS,S/AS01 malaria vaccine with or without a booster dose in infants and children in Africa: final results of a phase 3, individually randomised, controlled trial. Lancet. 2015 Jul 4;386(9988):31–45.

23. Asante KP, Adjei G, Enuameh Y, Owusu-Agyei S. RTS,S malaria vaccine development: progress and considerations for postapproval introduction. VDT. 2016 Jun;25.

24. Collins KA, Snaith R, Cottingham MG, Gilbert SC, Hill AVS. Enhancing protective immunity to malaria with a highly immunogenic virus-like particle vaccine. Sci Rep. 2017 Apr 19;7:46621.

25. Ballou WR, Rothbard J, Wirtz RA, Gordon DM, Williams JS, Gore RW, et al. Immunogenicity of synthetic peptides from circumsporozoite protein of Plasmodium falciparum. Science. 1985 May 24;228(4702):996–9.

26. Datoo MS, Natama MH, Somé A, Traoré O, Rouamba T, Bellamy D, et al. Efficacy of a low-dose candidate malaria vaccine, R21 in adjuvant Matrix-M, with seasonal administration to children in Burkina Faso: a randomised controlled trial. Lancet. 2021 May 15;397(10287):1809–18.

27. Datoo MM, Dicko A, Tinto H, Ouédraogo J-B, Hamaluba M, Olotu A, et al. A Phase III Randomised Controlled Trial Evaluating the Malaria Vaccine Candidate R21/Matrix-M^TM^ in African Children. 2023;

28. Neafsey DE, Juraska M, Bedford T, Benkeser D, Valim C, Griggs A, et al. Genetic diversity and protective efficacy of the RTS,S/AS01 malaria vaccine. N Engl J Med. 2015 Nov 19;373(21):2025–37.

29. Chaudhury S, MacGill RS, Early AM, Bolton JS, King CR, Locke E, et al. Breadth of humoral immune responses to the C-terminus of the circumsporozoite protein is associated with protective efficacy induced by the RTS,S malaria vaccine. Vaccine. 2021 Feb 5;39(6):968–75.

30. Pinzon-Ortiz C, Friedman J, Esko J, Sinnis P. The binding of the circumsporozoite protein to cell surface heparan sulfate proteoglycans is required for plasmodium sporozoite attachment to target cells. J Biol Chem. 2001 Jul 20;276(29):26784–91.

31. Rathore D, Sacci JB, de la Vega P, McCutchan TF. Binding and invasion of liver cells by Plasmodium falciparum sporozoites. Essential involvement of the amino terminus of circumsporozoite protein. J Biol Chem. 2002 Mar 1;277(9):7092–8.

32. Gandhi K, Thera MA, Coulibaly D, Traoré K, Guindo AB, Ouattara A, et al. Variation in the circumsporozoite protein of Plasmodium falciparum: vaccine development implications. PLoS ONE. 2014 Jul 3;9(7):e101783.

33. Gandhi K, Thera MA, Coulibaly D, Traoré K, Guindo AB, Doumbo OK, et al. Next generation sequencing to detect variation in the Plasmodium falciparum circumsporozoite protein. Am J Trop Med Hyg. 2012 May;86(5):775–81.

34. Egan, JE, Hoffman SL, Gordon DM. Humoral immune responses in volunteers immunized with irradiated Plasmodium falciparum sporozoites. Cover The American Journal of Tropical Medicine and Hygiene The American Journal of Tropical Medicine and Hygiene. 1993;

35. Plassmeyer ML, Reiter K, Shimp RL, Kotova S, Smith PD, Hurt DE, et al. Structure of the Plasmodium falciparum circumsporozoite protein, a leading malaria vaccine candidate. J Biol Chem. 2009 Sep 25;284(39):26951–63.

36. Barry AE, Schultz L, Buckee CO, Reeder JC. Contrasting population structures of the genes encoding ten leading vaccine-candidate antigens of the human malaria parasite, Plasmodium falciparum. PLoS ONE. 2009 Dec 30;4(12):e8497.

37. Zeeshan M, Alam MT, Vinayak S, Bora H, Tyagi RK, Alam MS, et al. Genetic variation in the Plasmodium falciparum circumsporozoite protein in India and its relevance to RTS,S malaria vaccine. PLoS ONE. 2012 Aug 17;7(8):e43430.

38. Lê HG, Kang J-M, Moe M, Jun H, Thái TL, Lee J, et al. Genetic polymorphism and natural selection of circumsporozoite surface protein in Plasmodium falciparum field isolates from Myanmar. Malar J. 2018 Oct 12;17(1):361.

39. Ishengoma D, Shayo A, Mandara C, Baraka V, Madebe R, Ngatunga, et al. The Role of Malaria Rapid DiagnosticTests in Screening of Patients to be Enrolled in Clinical Trials in Low Malaria Transmission Settings. iMedPub Journals. 2016 Mar 16;

40. MalariaGEN, Ahouidi A, Ali M, Almagro-garcia J, Amambua-ngwa A, Amaratunga C, et al. An open dataset of Plasmodium falciparum genome variation in 7,000 worldwide samples. Wellcome Open Research. 2021;1–22.

41. Ghansah A, Amenga-Etego L, Amambua-Ngwa A, Andagalu B, Apinjoh T, Bouyou-Akotet M, et al. Monitoring parasite diversity for malaria elimination in sub-Saharan Africa. Science. 2014 Sep 12;345(6202):1297–8.

42. Shayo A, Mandara CI, Shahada F, Buza J, Lemnge MM, Ishengoma DS. Therapeutic efficacy and safety of artemether-lumefantrine for the treatment of uncomplicated falciparum malaria in North-Eastern Tanzania. Malar J. 2014 Sep 20;13:376.

43. Mandara CI, Kavishe RA, Gesase S, Mghamba J, Ngadaya E, Mmbuji P, et al. High efficacy of artemether-lumefantrine and dihydroartemisinin-piperaquine for the treatment of uncomplicated falciparum malaria in Muheza and Kigoma Districts, Tanzania. Malar J. 2018 Jul 11;17(1):261.

44. Venkatesan M, Amaratunga C, Campino S, Auburn S, Koch O, Lim P, et al. Using CF11 cellulose columns to inexpensively and effectively remove human DNA from Plasmodium falciparum-infected whole blood samples. Malar J. 2012 Feb 10;11:41.

45. MalariaGEN. Pf7: an open dataset of Plasmodium falciparum genome. Wellcome Open Research. 2023;8(22).

46. MalariaGEN, Ahouidi A, Ali M, Almagro-Garcia J, Amambua-Ngwa A, Amaratunga C, et al. An open dataset of Plasmodium falciparum genome variation in 7,000 worldwide samples. Wellcome Open Res. 2021 Jul 13;6:42.

47. Jung Y, Han D. BWA-MEME: BWA-MEM emulated with a machine learning approach. BioRxiv. 2021 Sep 1;

48. Purcell S, Neale B, Todd-Brown K, Thomas L, Ferreira MAR, Bender D, et al. PLINK: a tool set for whole-genome association and population-based linkage analyses. Am J Hum Genet. 2007 Sep;81(3):559–75.

49. McKenna A, Hanna M, Banks E, Sivachenko A, Cibulskis K, Kernytsky A, et al. The Genome Analysis Toolkit: a MapReduce framework for analyzing next-generation DNA sequencing data. Genome Res. 2010 Sep;20(9):1297–303.

50. Manske M, Miotto O, Campino S, Auburn S, Almagro-Garcia J, Maslen G, et al. Analysis of *Plasmodium falciparum* diversity in natural infections by deep sequencing. Nature. 2012 Jul 19;487(7407):375–9.

51. Lee S, Harrison A, Tessier N, Tavul L, Miotto O. Assessing clonality in malaria parasites from massively parallel sequencing data. F1000Research. 2015;4(1043).

52. Chakraborty R, Danker-Hopfe H. Analysis of population structure: A comparative study of different estimators of wright’s fixation indices. Statistical methods in biological and medical sciences. Elsevier; 1991. p. 203–54.

53. Rozas J, Ferrer-Mata A, Sánchez-DelBarrio JC, Guirao-Rico S, Librado P, Ramos-Onsins SE, et al. Dnasp 6: DNA sequence polymorphism analysis of large data sets. Mol Biol Evol. 2017 Dec 1;34(12):3299–302.

54. Bandelt HJ, Forster P, Röhl A. Median-joining networks for inferring intraspecific phylogenies. Mol Biol Evol. 1999 Jan;16(1):37–48.

55. Danecek P, Auton A, Abecasis G, Albers CA, Banks E, DePristo MA, et al. The variant call format and VCFtools. Bioinformatics. 2011 Aug 1;27(15):2156–8.

56. Tajima F. Statistical method for testing the neutral mutation hypothesis by DNA polymorphism. Genetics. 1989 Nov;123(3):585–95.

57. Fu YX, Li WH. Statistical tests of neutrality of mutations. Genetics. 1993 Mar;133(3):693–709.

58. Tamura K, Stecher G, Kumar S. MEGA11: Molecular Evolutionary Genetics Analysis version 11. Mol Biol Evol. 2021 Jun 25;38(7):3022–7.

59. Thawer SG, Chacky F, Runge M, Reaves E, Mandike R, Lazaro S, et al. Sub-national stratification of malaria risk in mainland Tanzania: a simplified assembly of survey and routine data. Malar J. 2020 May 8;19(1):177.

60. MoHCDGEC. NATIONAL MALARIA STRATEGIC PLAN 2021-2025, TRANSITIONING TO MALARIA ELIMINATION IN PHASES. Ministry of Health, Community Development, Gender, Elderly and Childre. 2021;

61. WHO. WHO recommends R21/Matrix-M vaccine for malaria prevention in updated advice on immunization. WHO. 2023;

62. Aragam NR, Thayer KM, Nge N, Hoffman I, Martinson F, Kamwendo D, et al. Diversity of T cell epitopes in Plasmodium falciparum circumsporozoite protein likely due to protein-protein interactions. PLoS ONE. 2013 May 7;8(5):e62427.

63. Pringle JC, Wesolowski A, Berube S, Kobayashi T, Gebhardt ME, Mulenga M, et al. High Plasmodium falciparum genetic diversity and temporal stability despite control efforts in high transmission settings along the international border between Zambia and the Democratic Republic of the Congo. Malar J. 2019 Dec 4;18(1):400.

64. Amegashie EA, Amenga-Etego L, Adobor C, Ogoti P, Mbogo K, Amambua-Ngwa A, et al. Population genetic analysis of the Plasmodium falciparum circumsporozoite protein in two distinct ecological regions in Ghana. Malar J. 2020 Nov 27;19(1):437.

65. He Z-Q, Zhang Q-Q, Wang D, Hu Y-B, Zhou R-M, Qian D, et al. Genetic polymorphism of circumsporozoite protein of Plasmodium falciparum among Chinese migrant workers returning from Africa to Henan Province. Malar J. 2022 Aug 27;21(1):248.

66. Duffy CW, Ba H, Assefa S, Ahouidi AD, Deh YB, Tandia A, et al. Population genetic structure and adaptation of malaria parasites on the edge of endemic distribution. Mol Ecol. 2017 Jun;26(11):2880–94.

67. Amambua-Ngwa A, Tetteh KKA, Manske M, Gomez-Escobar N, Stewart LB, Deerhake ME, et al. Population genomic scan for candidate signatures of balancing selection to guide antigen characterization in malaria parasites. PLoS Genet. 2012 Nov 1;8(11):e1002992.

68. Pringle JC, Carpi G, Almagro-Garcia J, Zhu SJ, Kobayashi T, Mulenga M, et al. RTS,S/AS01 malaria vaccine mismatch observed among Plasmodium falciparum isolates from southern and central Africa and globally. Sci Rep. 2018 Apr 26;8(1):6622.

69. Bailey JA, Mvalo T, Aragam N, Weiser M, Congdon S, Kamwendo D, et al. Use of massively parallel pyrosequencing to evaluate the diversity of and selection on Plasmodium falciparum csp T-cell epitopes in Lilongwe, Malawi. J Infect Dis. 2012 Aug 15;206(4):580–7.

70. Polley SD, Conway DJ. Strong diversifying selection on domains of the Plasmodium falciparum apical membrane antigen 1 gene. Genetics. 2001 Aug;158(4):1505–12.

71. Nirmolia T, Ahmed MA, Sathishkumar V, Sarma NP, Bhattacharyya DR, Mohapatra PK, et al. Genetic diversity of Plasmodium falciparum AMA-1 antigen from the Northeast Indian state of Tripura and comparison with global sequences: implications for vaccine development. Malar J. 2022 Feb 22;21(1):62.

72. Ajibola O, Diop MF, Ghansah A, Amenga-Etego L, Golassa L, Apinjoh T, et al. In silico characterisation of putative *Plasmodium falciparum* vaccine candidates in African malaria populations. Sci Rep. 2021 Aug 10;11(1):16215.

73. Chenet SM, Branch OH, Escalante AA, Lucas CM, Bacon DJ. Genetic diversity of vaccine candidate antigens in Plasmodium falciparum isolates from the Amazon basin of Peru. Malar J. 2008 May 27;7:93.

74. Reeder JC, Wapling J, Mueller I, Siba PM, Barry AE. Population genetic analysis of the Plasmodium falciparum 6-cys protein Pf38 in Papua New Guinea reveals domain-specific balancing selection. Malar J. 2011 May 14;10:126.

75. DHS. Malaria Indicator Survey 2017 [Internet]. 2018 [cited 2024 Jan 15]. Available from: https://dhsprogram.com/pubs/pdf/MIS31/MIS31.pdf

